# The Bidirectional Relationship Between Coronary Artery Disease and Epilepsy: Cross-sectional and Prospective Cohort Studies from the UK Biobank

**DOI:** 10.1101/2025.08.01.25332722

**Authors:** Li Zhang, Weifang Liu, Yilu Xue, Liwen Wang, Cong Gong, Zhi-Gang She, Hongliang Li

**Affiliations:** Department of Cardiology, Renmin Hospital of Wuhan University, Wuhan, Hubei Province, China; State Key Laboratory of New Drug Discovery and Development for Major Diseases, Gannan Medical University; Gannan Innovation and Translational Medicine Research Institute, Gannan Medical University, Ganzhou, China

**Keywords:** Bidirectional relationship, Epilepsy, Coronary artery disease, Polygenic risk score

## Abstract

**Background and aims:** Coronary artery disease (CAD) is the leading cause of morbidity and mortality globally, while epilepsy is a common neurological disorder. Previous studies have compared the cardiovascular comorbidity burden in patients with epilepsy and their risk of cardiovascular mortality during follow-up. However, the bidirectional relationship between CAD and epilepsy remains unexplored. This study comprehensively assessed the association between CAD and epilepsy using cross-sectional and cohort designs in the UK Biobank, and further investigated whether genetic susceptibility influences this link.

**Methods:** This population-based study utilized UK Biobank data to investigate the relationship between epilepsy and CAD. A cross-sectional analysis of 502,359 participants was conducted using logistic regression to estimate odds ratios (ORs). In cohort 1 (n = 496,921 without baseline epilepsy), stratified Cox models assessed the risk of incident epilepsy by CAD status. In cohort 2 (n = 475,130 without baseline CAD), the risk of incident CAD by epilepsy status was similarly evaluated. Polygenic risk scores (PRS) for both CAD and epilepsy were constructed to explore genetic susceptibility.

**Results:** The median age at baseline was 58.0 years, and 45.60% were male. During a median follow-up of 13.82 years for epilepsy and 13.73 years for CAD, 3,590 participants developed epilepsy and 39,223 developed CAD. In cross-sectional analysis, CAD was significantly associated with epilepsy. In Cohort 1, CAD was associated with higher epilepsy risk (HR: 1.29; 95% CI: 1.15–1.45; P < 0.001), and in Cohort 2, epilepsy was associated with higher CAD risk (HR: 1.34; 95% CI: 1.18–1.52; P < 0.001). These associations persisted across subgroups by age, sex, and BMI. Interestingly, CAD was linked to incident epilepsy only in participants with low genetic risk, while epilepsy predicted CAD only in those with high genetic susceptibility.

**Conclusions:** Our findings indicate a bidirectional association between CAD and epilepsy, while also exploring the heterogeneity of this association across different subpopulations and its potential modification by genetic susceptibility. These results underscore the need for further studies to elucidate the underlying mechanisms and clinical implications of this association.

## Introduction

CAD is the leading cause of morbidity and mortality globally. ^1–3^ According to the Global Burden of Disease (GBD) study, CAD was diagnosed in an estimated 197 million individuals worldwide in 2019, causing 9.14 million deaths and accounting for 49.2% of all CVD-related fatalities.^1,9^ While the established risk factors for CAD have been extensively studied, the potential contributions of nervous system disorders-particularly epilepsy-have not been fully elucidated.^4–7^

Epilepsy is a prevalent neurological disorder, impacting more than 70 million individuals globally.^13–17^ Beyond its effects on physical health, epilepsy also imposes significant psychological, social, and economic burdens.^18–20^ Moreover, studies suggest that epilepsy may elevate the risk of CAD in affected individuals by promoting the release of catecholamines and glucocorticoids, while CAD-related hypoperfusion or myocardial injury may exacerbate the frequency and severity of seizures, creating a vicious cycle.^46–49^

Despite these insights, the interplay between epilepsy and CAD remains under explored. To date, only two small-scale cross-sectional studies in the United States have demonstrated a significantly higher prevalence of heart disease in individuals with epilepsy, as well as an increased prevalence of epilepsy in patients with heart disease.^24–25^ Additionally, several European studies have investigated the cardiovascular risks and mortality associated with epilepsy, indicating that individuals with epilepsy have a higher prevalence of cardiovascular disease and increased cardiovascular mortality compared to those without epilepsy.^26–27^ Furthermore, patients with comorbid epilepsy and cardiovascular disease appear to have an even greater risk of death.^28^ These studies suggest that epilepsy may be a risk factor for cardiovascular mortality. Nevertheless, no studies have specifically examined the role of CAD in the onset of epilepsy, nor have they explored the potential contribution of epilepsy to the development of CAD. Moreover, the possible influence of genetic susceptibility on the relationship between these two conditions remains unexplored.

In this study, we conducted a comprehensive analysis using data from the UK Biobank to investigate the bidirectional relationship between CAD and epilepsy. First, we assessed the cross-sectional associations between CAD and epilepsy. Subsequently, we performed prospective cohort analyses to evaluate the association between baseline CAD and subsequent epilepsy and the association between baseline epilepsy and subsequent CAD. We further explored subgroup differences based on demographic and clinical characteristics and investigated the moderating effect of genetic predisposition. This research not only advances our understanding of the shared pathophysiological mechanisms of CAD and epilepsy but also holds promise for informing integrated management strategies and personalized treatment approaches.

## Materials and methods

### Data source and study design

The UK Biobank is a large population-based prospective study that recruited over 500,000 participants aged 40-69 years in the UK between 2006 and 2010.^29^ The researchers gathered information on the participants’ sociodemographic characteristics, lifestyle choices, medical history, and physical measurements at recruitment. Ethical approval for the study was obtained from the National Health Service (NHS) North West Research Ethics Committee, and all participants signed an informed consent form. The study was conducted under UK Biobank application number 77195. More comprehensive information is available at https://www.ukbiobank.ac.uk/.

In the present study, we synthesized evidence from one cross-sectional study and two prospective cohort studies. First, among 502,412 participants in the UK Biobank, we excluded 53 participants who withdrew informed consent, leaving 502,359 participants included in the cross-sectional analysis to explore the associations between CAD and epilepsy. Subsequently, we conducted two prospective cohort studies. For Cohort 1, we excluded 5,438 participants with a diagnosis of epilepsy at baseline, leaving 496,921 participants to investigate the association between baseline CAD and the risk of developing epilepsy. For Cohort 2, we excluded 27,229 participants with CAD at baseline, leaving 475,130 participants to examine the relationship between epilepsy and the incidence of new-onset CAD. The complete participant selection process is illustrated in **Figure 1**.

**Figure 1.**
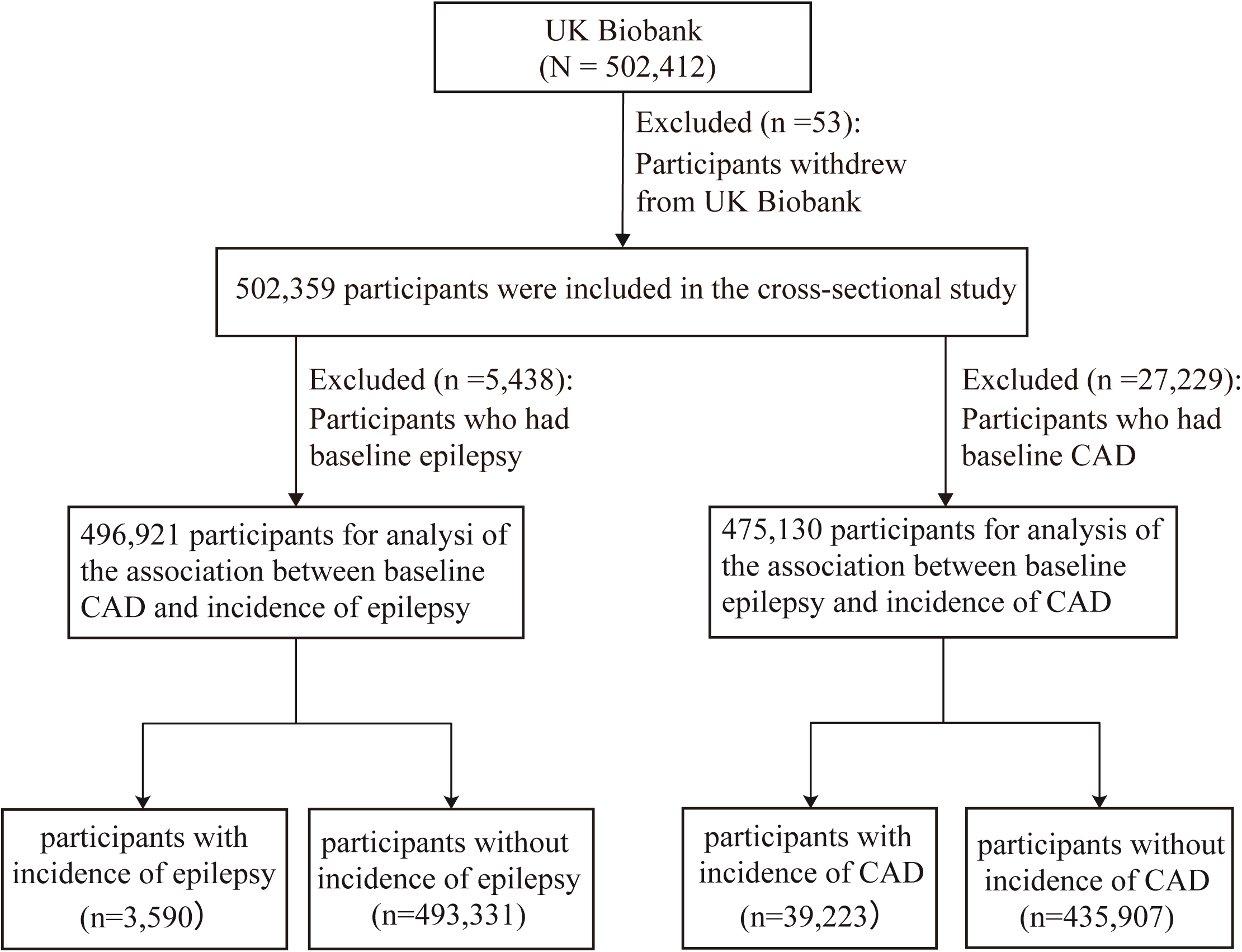
Flow chart of participants. Abbreviation: CAD, coronary artery disease.

### Ascertainment of CAD and epilepsy

Cases of CAD and epilepsy were identified from health-related records of the International Classification of Diseases, 10th Edition (ICD-10), coded from self-reported information, primary care data, death registration records, and hospitalization data. The diagnostic codes for CAD are ICD-10 I20-I25, and the diagnostic codes for epilepsy are ICD-10 G40 and G41. Follow-up commenced upon participants’ enrollment and persisted until the date of diagnosis, loss of follow-up, mortality, or conclusion of the study (January 2, 2023)—whichever came first.

### Data collection

We collected covariates based on a variety of data sources, including medical history, physical measurements, touch-screen questionnaires, and laboratory indicators. These covariates included age (continuous), sex (male/female), body mass index (BMI) (continuous), ethnicity (White, Asian or Asian British, Black or Black British, Chinese, Mixed, Other), smoking status (current/previous/never), alcohol consumption (current/previous/never), physical activity level (high/moderate/low), education level (college/university degree, A/AS levels, NVQ, HND, HNC, other professional qualifications, O levels/GCSEs or equivalent, none), and Townsend deprivation index (an area-based measure of socioeconomic status that incorporates unemployment, lack of car ownership, non-home ownership, and household overcrowding; higher scores indicate greater material deprivation) (continuous). Additionally, we considered laboratory and clinical indicators including total triglycerides (continuous), total cholesterol (continuous), HDL cholesterol (HDL-C) (continuous), LDL cholesterol (LDL-C) (continuous), systolic blood pressure (SBP) (continuous), diastolic blood pressure (DBP) (continuous), glucose (GLU) (continuous), and C-reactive protein (CRP) (continuous). Furthermore, comorbidities and therapeutic medications relevant to CAD and epilepsy, such as hypertension, hyperlipidemia, diabetes, depression, cerebrovascular disease, hyperthyroidism, central nervous system infections, lipid-lowering drugs, antihypertensive medications, and insulin, were also considered.

### Polygenic risk score

The genotypic data, quality control procedures, polygenic risk score (PRS) construction, and evaluation for the UK Biobank have been described in previous reports.^30^ In Cohort 1, we excluded participants who failed genetic data quality control (N=14,999); in Cohort 2, we excluded those with missing genetic data (N=10,328). The PRS for CAD was obtained from the ‘Standard PRS’ constructed by the UK Biobank with field ID: 26227 [Standard PRS for coronary artery disease (CAD)].^31^ For the PRS of epilepsy, we acquired the imputed genotype data from UK Biobank, and extracted epilepsy-related single nucleotide polymorphisms (SNPs) from published genome-wide association studies.^32^ Each SNP was coded as 0, 1, or 2 depending on the number of risk alleles, and we calculated the PRS for each participant using a weighting method: PRS = β_1_*SNP_1_ + β_2_*SNP_2_ +… + β_n_*SNP_n_. A higher PRS indicates greater genetic susceptibility to CAD or epilepsy.^33–36^ Subsequently, we divided participants into a low genetic risk group and a high genetic risk group based on their median PRS.

### Statistical analyses

Baseline characteristics of participants were presented as the median with interquartile range (IQR) for continuous variables and as frequencies and percentages for categorical variables. The difference between groups was tested by the Mann-Whitney U test. Categorical variables were compared using the chi-square test or Fisher’s exact test. The MICE package in R was utilized to impute the missing data on the variables through the chained equations method. The comprehensive missing rates for the covariates utilized in this study are presented in **Supplemental Table 1**.

In the cross-sectional study, logistics regression was used to evaluate the associations between CAD and epilepsy, with results expressed as odds ratio (OR) value and 95% confidence interval (CI). In the cohort study, stratified Cox proportional hazards regression was used to evaluate the relationship between baseline CAD and subsequent epilepsy, as well as between epilepsy and new-onset CAD. The results were expressed as hazard ratios (HR) and 95% CI. The proportional hazards assumption for the stratified Cox model was tested using the Schoenfeld residuals method, which confirmed that the assumption was satisfied. Both the logistic regression models and stratified Cox proportional regression models employed three adjustment models. Model 1 adjusted for age, sex, and ethnicity. Model 2 further adjusted for BMI, smoking status, alcohol consumption, physical activity, education level, and the Townsend Deprivation Index. Building upon Model 2, Model 3 additionally adjusted for SBP, laboratory test measures (total triglyceride, total cholesterol, HDL-C, LDL-C, GLU, and CRP), and comorbidity/medication history. Comorbidities included hypertension, hyperlipidemia, diabetes mellitus, depression, cerebrovascular disease, hyperthyroidism, CNS infections (Cohort 1 only), and CVD family history (Cohort 2 only). Therapeutic medications included lipid-lowering drugs, antihypertensive medications, and insulin.

To investigate potential variations in the bidirectional relationship between CAD and epilepsy across different subgroups, we conducted stratified analyses based on the following factors: age (≤60 years vs. >60 years), sex (male vs. female), BMI (≤25 vs. >25 kg/m²), ethnicity (White vs. non-White), smoking status (current, previous, or never), alcohol consumption status (current, previous, or never), frequency of physical activity (high, moderate, or low), and comorbidity status (none vs. at least one comorbidity). To further evaluate the role of genetic susceptibility in the bidirectional association between CAD and epilepsy, subgroup analyses stratified by genetic risk were conducted. Furthermore, polygenic risk scores (PRS) were incorporated into the primary analysis to account for potential confounding effects of genetic factors.

We conducted several sensitivity analyses to test the robustness of the main findings of this study: (1) We excluded participants without major covariates in the cross-sectional and cohort studies and repeated the main analysis. (2) We excluded participants in the longitudinal cohort with follow-up durations of less than two years and repeated the main analysis. (3) We extended the proportional risk model using a competing risk model, with death from other causes treated as a competing risk, to evaluate the stability of the findings. (4) We used a mixed-effects model in the longitudinal cohort to adjust for potential effects of the UK Biobank Assessment Centre and repeated the main analysis.

All statistical analyses were performed by R-4.0.0 (R Foundation for Statistical Computing, Vienna, Austria). *P* values <0.05 were considered statistically significant.

## Results

### Baseline Characteristics of Individuals in the Cross-Sectional Study

**Table 1** presents the baseline characteristics of 502,359 participants from the cross-sectional study, including 27,229 (5.42%) with CAD and 5,438 (1.08%) with epilepsy. The median age of all participants was 58.0 years (IQR, 50.0, 63.0), with 45.6% being male, 94.59% of White ethnicity, and a median BMI of 26.74 kg/m^2^ (IQR, 24.14, 29.91). Compared with the non-CAD and non-epilepsy groups, participants in the CAD and epilepsy groups had higher BMI, lower DBP, higher CRP, higher smoking frequency, lower drinking frequency, lower exercise frequency, lower education level, and lower Townsend deprivation index (all *P* < 0.001). These groups also had a higher prevalence of baseline comorbidities (all *P* < 0.001). Furthermore, they were more likely to use lipid-lowering drugs, blood pressure medications, and insulin (all *P* < 0.001). Notably, the prevalence of epilepsy was significantly higher in the CAD group than in the non-CAD group (1.78% vs 1.04%, *P* < 0.001). Similarly, the prevalence of CAD was higher in individuals with epilepsy compared to those without epilepsy (8.92% vs 5.38%, *P* < 0.001).

**Table 1.**
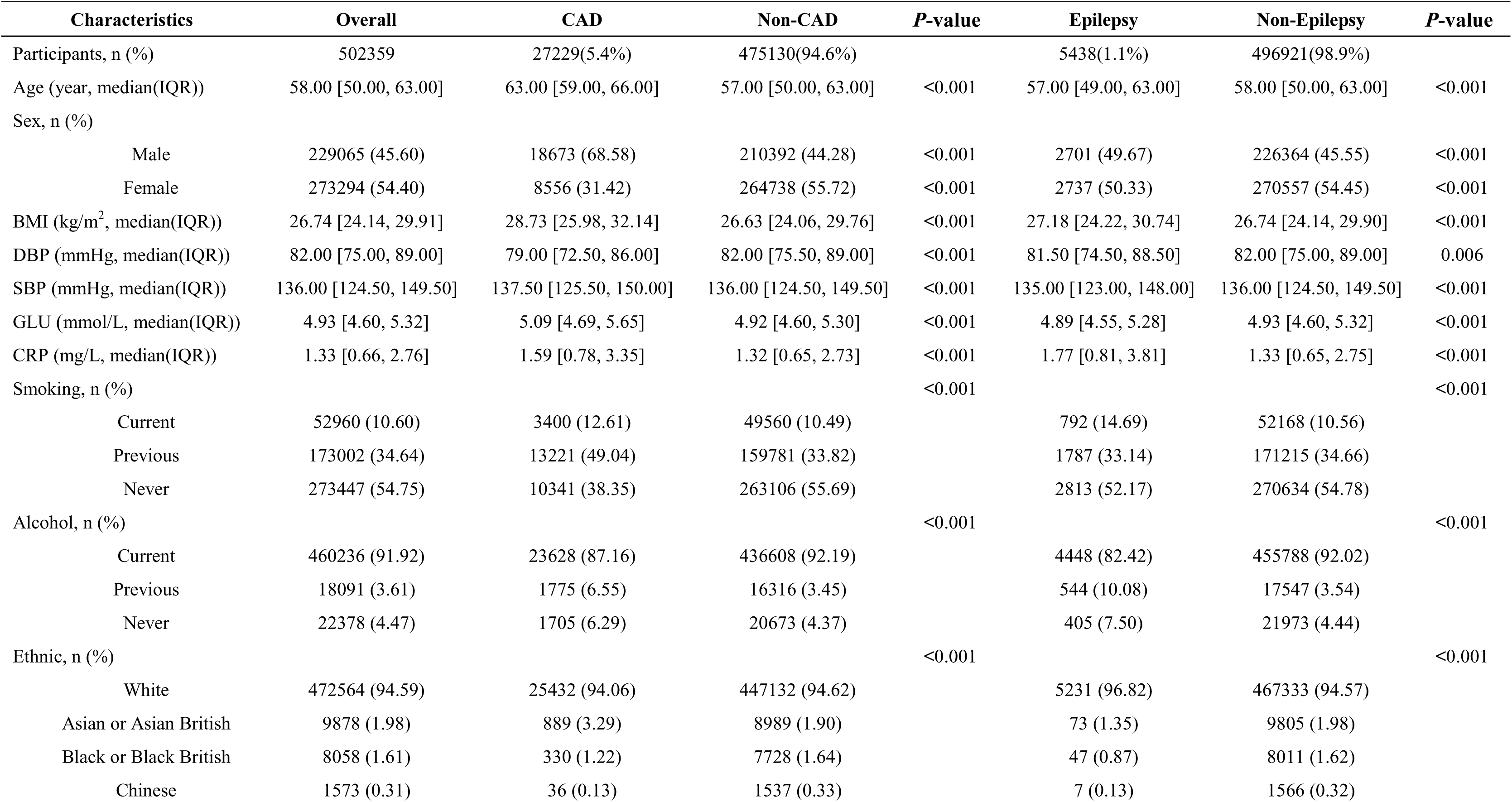
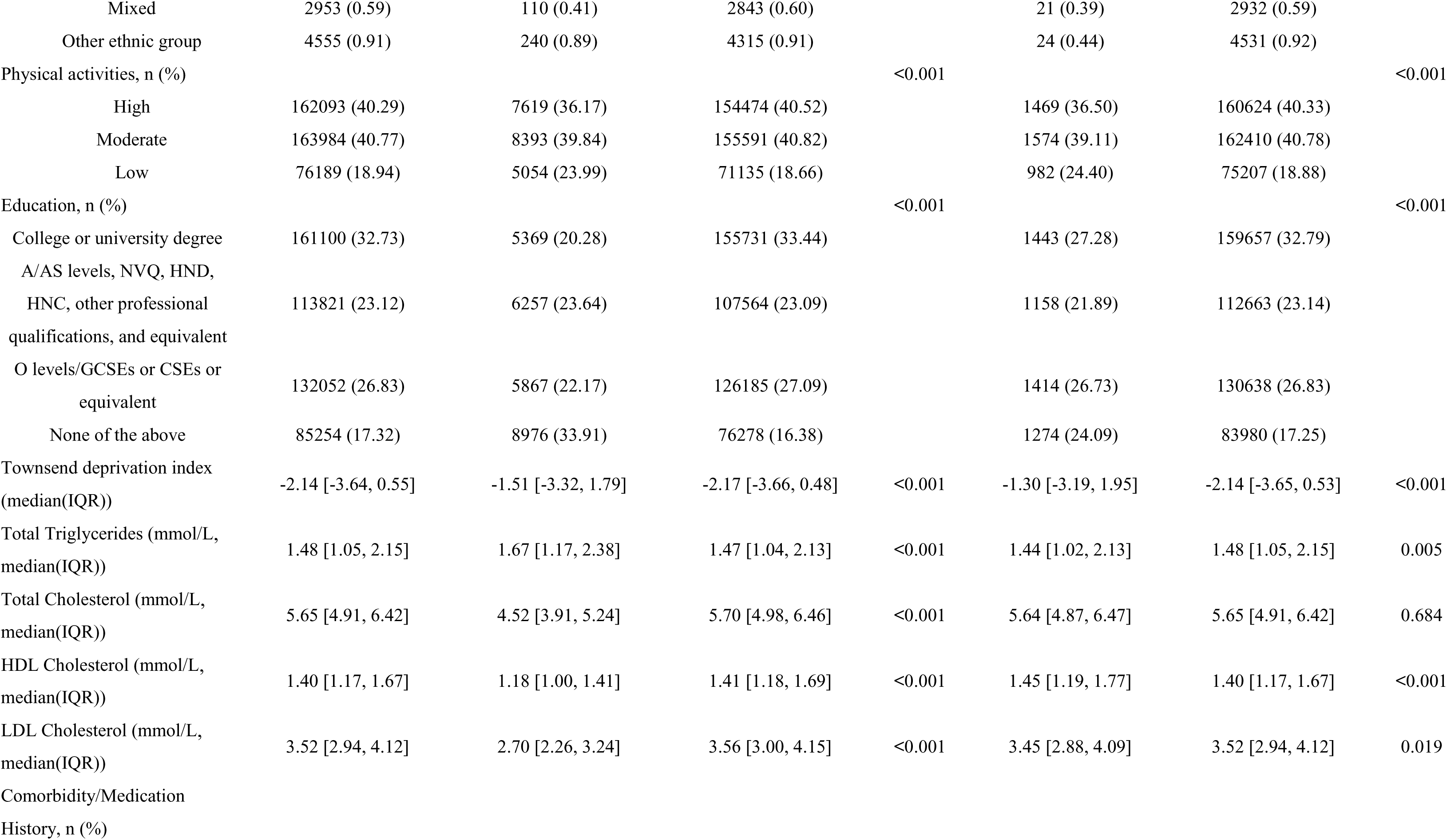
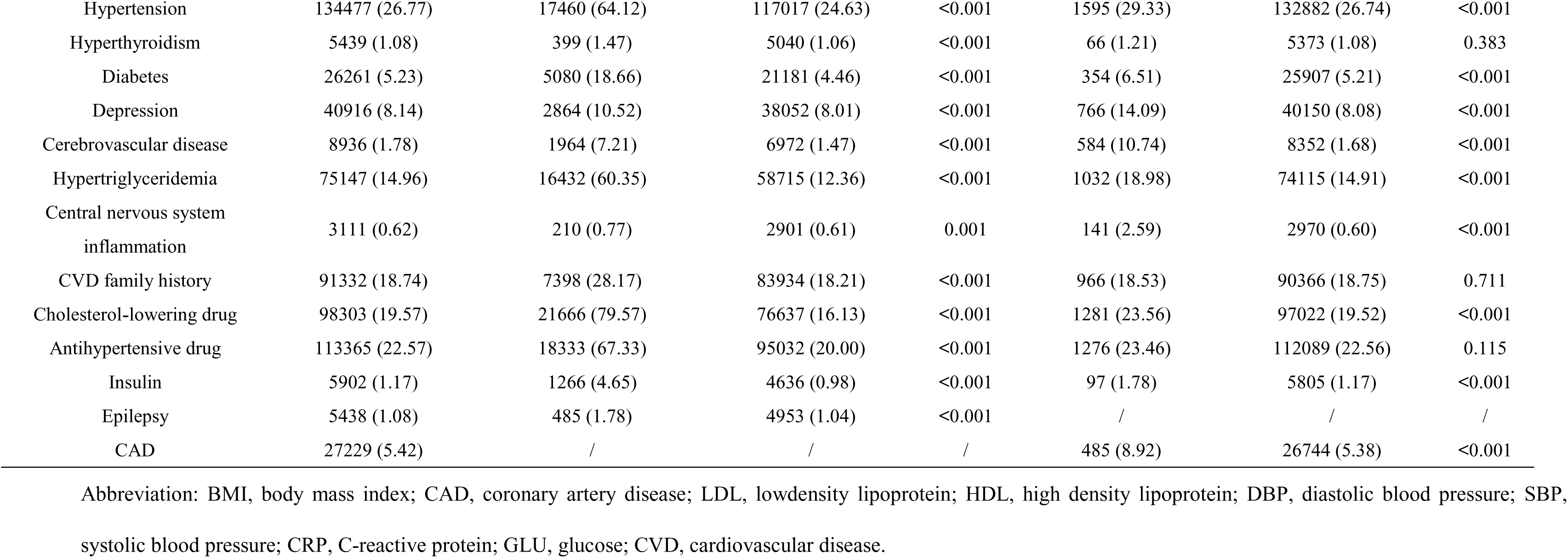
Baseline characteristics of 502,359 participants stratified by CAD and epilepsy in the Cross-Sectional Study.

| Characteristics | Overall | CAD | Non-CAD | P-value | Epilepsy | Non-Epilepsy | P-value |
| --- | --- | --- | --- | --- | --- | --- | --- |
| Participants, n (%) | 502359 | 27229(5.4%) | 475130(94.6%) |  | 5438(1.1%) | 496921(98.9%) |  |
| Age (year, median(IQR)) | 58.00 [50.00, 63.00] | 63.00 [59.00, 66.00] | 57.00 [50.00, 63.00] | <0.001 | 57.00 [49.00, 63.00] | 58.00 [50.00, 63.00] | <0.001 |
| Sex, n (%) |  |  |  |  |  |  |  |
| Male | 229065 (45.60) | 18673 (68.58) | 210392 (44.28) | <0.001 | 2701 (49.67) | 226364 (45.55) | <0.001 |
| Female | 273294 (54.40) | 8556 (31.42) | 264738 (55.72) | <0.001 | 2737 (50.33) | 270557 (54.45) | <0.001 |
| BMI (kg/m<sup>2</sup>, median(IQR)) | 26.74 [24.14, 29.91] | 28.73 [25.98, 32.14] | 26.63 [24.06, 29.76] | <0.001 | 27.18 [24.22, 30.74] | 26.74 [24.14, 29.90] | <0.001 |
| DBP (mmHg, median(IQR)) | 82.00 [75.00, 89.00] | 79.00 [72.50, 86.00] | 82.00 [75.50, 89.00] | <0.001 | 81.50 [74.50, 88.50] | 82.00 [75.00, 89.00] | 0.006 |
| SBP (mmHg, median(IQR)) | 136.00 [124.50, 149.50] | 137.50 [125.50, 150.00] | 136.00 [124.50, 149.50] | <0.001 | 135.00 [123.00, 148.00] | 136.00 [124.50, 149.50] | <0.001 |
| GLU (mmol/L, median(IQR)) | 4.93 [4.60, 5.32] | 5.09 [4.69, 5.65] | 4.92 [4.60, 5.30] | <0.001 | 4.89 [4.55, 5.28] | 4.93 [4.60, 5.32] | <0.001 |
| CRP (mg/L, median(IQR)) | 1.33 [0.66, 2.76] | 1.59 [0.78, 3.35] | 1.32 [0.65, 2.73] | <0.001 | 1.77 [0.81, 3.81] | 1.33 [0.65, 2.75] | <0.001 |
| Smoking, n (%) |  |  |  | <0.001 |  |  | <0.001 |
| Current | 52960 (10.60) | 3400 (12.61) | 49560 (10.49) |  | 792 (14.69) | 52168 (10.56) |  |
| Previous | 173002 (34.64) | 13221 (49.04) | 159781 (33.82) |  | 1787 (33.14) | 171215 (34.66) |  |
| Never | 273447 (54.75) | 10341 (38.35) | 263106 (55.69) |  | 2813 (52.17) | 270634 (54.78) |  |
| Alcohol, n (%) |  |  |  | <0.001 |  |  | <0.001 |
| Current | 460236 (91.92) | 23628 (87.16) | 436608 (92.19) |  | 4448 (82.42) | 455788 (92.02) |  |
| Previous | 18091 (3.61) | 1775 (6.55) | 16316 (3.45) |  | 544 (10.08) | 17547 (3.54) |  |
| Never | 22378 (4.47) | 1705 (6.29) | 20673 (4.37) |  | 405 (7.50) | 21973 (4.44) |  |
| Ethnic, n (%) |  |  |  | <0.001 |  |  | <0.001 |
| White | 472564 (94.59) | 25432 (94.06) | 447132 (94.62) |  | 5231 (96.82) | 467333 (94.57) |  |
| Asian or Asian British | 9878 (1.98) | 889 (3.29) | 8989 (1.90) |  | 73 (1.35) | 9805 (1.98) |  |
| Black or Black British | 8058 (1.61) | 330 (1.22) | 7728 (1.64) |  | 47 (0.87) | 8011 (1.62) |  |
| Chinese | 1573 (0.31) | 36 (0.13) | 1537 (0.33) |  | 7 (0.13) | 1566 (0.32) |  |
| Mixed | 2953 (0.59) | 110 (0.41) | 2843 (0.60) |  | 21 (0.39) | 2932 (0.59) |  |
| Other ethnic group | 4555 (0.91) | 240 (0.89) | 4315 (0.91) |  | 24 (0.44) | 4531 (0.92) |  |
| Physical activities, n (%) |  |  |  | <0.001 |  |  | <0.001 |
| High | 162093 (40.29) | 7619 (36.17) | 154474 (40.52) |  | 1469 (36.50) | 160624 (40.33) |  |
| Moderate | 163984 (40.77) | 8393 (39.84) | 155591 (40.82) |  | 1574 (39.11) | 162410 (40.78) |  |
| Low | 76189 (18.94) | 5054 (23.99) | 71135 (18.66) |  | 982 (24.40) | 75207 (18.88) |  |
| Education, n (%) |  |  |  | <0.001 |  |  | <0.001 |
| College or university degree | 161100 (32.73) | 5369 (20.28) | 155731 (33.44) |  | 1443 (27.28) | 159657 (32.79) |  |
| A/AS levels, NVQ, HND, HNC, other professional qualifications, and equivalent | 113821 (23.12) | 6257 (23.64) | 107564 (23.09) |  | 1158 (21.89) | 112663 (23.14) |  |
| O levels/GCSEs or CSEs or equivalent | 132052 (26.83) | 5867 (22.17) | 126185 (27.09) |  | 1414 (26.73) | 130638 (26.83) |  |
| None of the above | 85254 (17.32) | 8976 (33.91) | 76278 (16.38) |  | 1274 (24.09) | 83980 (17.25) |  |
| Townsend deprivation index (median(IQR)) | -2.14 [-3.64, 0.55] | -1.51 [-3.32, 1.79] | -2.17 [-3.66, 0.48] | <0.001 | -1.30 [-3.19, 1.95] | -2.14 [-3.65, 0.53] | <0.001 |
| Total Triglycerides (mmol/L, median(IQR)) | 1.48 [1.05, 2.15] | 1.67 [1.17, 2.38] | 1.47 [1.04, 2.13] | <0.001 | 1.44 [1.02, 2.13] | 1.48 [1.05, 2.15] | 0.005 |
| Total Cholesterol (mmol/L, median(IQR)) | 5.65 [4.91, 6.42] | 4.52 [3.91, 5.24] | 5.70 [4.98, 6.46] | <0.001 | 5.64 [4.87, 6.47] | 5.65 [4.91, 6.42] | 0.684 |
| HDL Cholesterol (mmol/L, median(IQR)) | 1.40 [1.17, 1.67] | 1.18 [1.00, 1.41] | 1.41 [1.18, 1.69] | <0.001 | 1.45 [1.19, 1.77] | 1.40 [1.17, 1.67] | <0.001 |
| LDL Cholesterol (mmol/L, median(IQR)) | 3.52 [2.94, 4.12] | 2.70 [2.26, 3.24] | 3.56 [3.00, 4.15] | <0.001 | 3.45 [2.88, 4.09] | 3.52 [2.94, 4.12] | 0.019 |
| Comorbidity/Medication History, n (%) |  |  |  |  |  |  |  |
| Hypertension | 134477 (26.77) | 17460 (64.12) | 117017 (24.63) | <0.001 | 1595 (29.33) | 132882 (26.74) | <0.001 |
| Hyperthyroidism | 5439 (1.08) | 399 (1.47) | 5040 (1.06) | <0.001 | 66 (1.21) | 5373 (1.08) | 0.383 |
| Diabetes | 26261 (5.23) | 5080 (18.66) | 21181 (4.46) | <0.001 | 354 (6.51) | 25907 (5.21) | <0.001 |
| Depression | 40916 (8.14) | 2864 (10.52) | 38052 (8.01) | <0.001 | 766 (14.09) | 40150 (8.08) | <0.001 |
| Cerebrovascular disease | 8936 (1.78) | 1964 (7.21) | 6972 (1.47) | <0.001 | 584 (10.74) | 8352 (1.68) | <0.001 |
| Hypertriglyceridemia | 75147 (14.96) | 16432 (60.35) | 58715 (12.36) | <0.001 | 1032 (18.98) | 74115 (14.91) | <0.001 |
| Central nervous system<br>inflammation | 3111 (0.62) | 210 (0.77) | 2901 (0.61) | 0.001 | 141 (2.59) | 2970 (0.60) | <0.001 |
| CVD family history | 91332 (18.74) | 7398 (28.17) | 83934 (18.21) | <0.001 | 966 (18.53) | 90366 (18.75) | 0.711 |
| Cholesterol-lowering drug | 98303 (19.57) | 21666 (79.57) | 76637 (16.13) | <0.001 | 1281 (23.56) | 97022 (19.52) | <0.001 |
| Antihypertensive drug | 113365 (22.57) | 18333 (67.33) | 95032 (20.00) | <0.001 | 1276 (23.46) | 112089 (22.56) | 0.115 |
| Insulin | 5902 (1.17) | 1266 (4.65) | 4636 (0.98) | <0.001 | 97 (1.78) | 5805 (1.17) | <0.001 |
| Epilepsy | 5438 (1.08) | 485 (1.78) | 4953 (1.04) | <0.001 | / | / | / |
| CAD | 27229 (5.42) | / | / | / | 485 (8.92) | 26744 (5.38) | <0.001 |

Abbreviation: BMI, body mass index; CAD, coronary artery disease; LDL, lowdensity lipoprotein; HDL, high density lipoprotein; DBP, diastolic blood pressure; SBP, systolic blood pressure; CRP, C-reactive protein; GLU, glucose; CVD, cardiovascular disease.

### Association of CAD and Epilepsy in the Cross-sectional Analysis

The risks associated with the presence of epilepsy in individuals with CAD and the presence of CAD in individuals with epilepsy are presented in **Table 2(A)** and **Table 2(B)**. Compared with the individuals without CAD, a significantly higher risk of epilepsy was found in individuals with CAD (after multivariate adjustment, OR, 1.25; 95% CI, 1.10-1.43, *P* < 0.001). Similarly, individuals with epilepsy had a significantly increased risk of CAD compared to those without epilepsy (after multivariate adjustment, OR, 1.45; 95% CI, 1.29-1.62, *P* < 0.001).

**Table 2.** Association Between CAD and Epilepsy in the Cross-Sectional Analysis.

**(A)**
| Groups | Model 1 |  | Model 2 |  | Model 3 |  |
| --- | --- | --- | --- | --- | --- | --- |
|  | OR (95% CI) | P-value | OR (95% CI) | P-value | OR (95% CI) | P-value |
| Non-CAD | 1 (reference) | - | 1 (reference) | - | 1 (reference) | - |
| CAD | 1.79 (1.62-1.97) | < 0.001 | 1.42 (1.26-1.60) | < 0.001 | 1.25 (1.10-1.43) | < 0.001 |

**(B)**
| Groups | Model 1 |  | Model 2 |  | Model 3 |  |
| --- | --- | --- | --- | --- | --- | --- |
|  | OR (95% CI) | P-value | OR (95% CI) | P-value | OR (95% CI) | P-value |
| Non-Epilepsy | 1 (reference) | - | 1 (reference) | - | 1 (reference) | - |
| Epilepsy | 1.79 (1.62-1.97) | < 0.001 | 1.47 (1.28-1.68) | < 0.001 | 1.45 (1.29-1.62) | < 0.001 |

Model 1: adjusted for age, sex, and ethnicity.
Model 2: adjusted for age, sex, BMI, ethnicity, alcohol drinking status, smoking status, physical activity, education, UK Biobank assessment centre, and Townsend deprivation index.
Model 3: adjusted for age, sex, BMI, ethnicity, alcohol drinking status, smoking status, physical activity, education, UK Biobank assessment centre, Townsend deprivation index, total cholesterol, LDL cholesterol, HDL cholesterol, total triglycerides, GLU, SBP, CRP, and comorbidity/medication history (hypertension, hyperthyroidism, diabetes, depression, cerebrovascular disease, hypertriglyceridemia, central nervous system inflammation, CVD family history, cholesterol-lowering drug, antihypertensive drug, insulin).
Abbreviation: CAD, coronary artery disease; BMI, body mass index; LDL, low-density lipoprotein; HDL, high-density lipoprotein; GLU, glucose; SBP, systolic blood pressure; CRP, C-reactive protein.

### Baseline Characteristics of Individuals in the Prospective Cohort Study

We subsequently conducted two longitudinal cohort studies to examine the temporal relationship between CAD and epilepsy. In cohort 1, we investigated the association between baseline CAD and subsequent epilepsy, using the non-CAD group as the control. In cohort 2, we explored the relationship between baseline epilepsy and new-onset CAD, with the non-epilepsy group as the control.

In Cohort 1, **Supplemental Table 2** presents the baseline characteristics of 496,921 participants, consisting of 26,744 (5.4%) with CAD and 470,177 (94.6%) without CAD. The median age was 58.0 years (IQR, 50.0–63.0), and 226,364 (45.55%) of the participants were male. Compared to those without baseline CAD, individuals with baseline CAD were more likely to be older, male, have a higher BMI, smoke frequently, be less educated, engage in less physical activity, and have higher baseline SBP (all *P* < 0.001). They also had a higher Townsend deprivation index, elevated total triglycerides, and lower total cholesterol, HDL-C, and LDL-C levels (all *P* < 0.001). Additionally, they used more lipid-lowering medications, antihypertensive drugs, insulin, and exhibited a higher prevalence of comorbidities (all *P* < 0.001).

In cohort 2, the baseline information of 475,130 participants involved in the analysis of the associations between epilepsy and the incidence of CAD is shown in **Supplementary Table 3**. Including 4,953 (1.0%) with epilepsy and 470,177 (99.0%) without epilepsy. The median age was 57.0 years (IQR, 50.0–63.0), with 44.3% of participants being male. In contrast to non-epilepsy participants, those with epilepsy exhibited a higher likelihood of being younger, male, having a higher BMI, being more frequent smokers, being white in ethnicity, being less educated, being less physically active, and scoring higher on the Townsend deprivation index (all *P* < 0.001). Additionally, individuals with epilepsy had elevated baseline CRP levels, greater use of lipid-lowering and antihypertensive medications, and insulin, as well as a higher prevalence of comorbidities (all *P* < 0.001).

### Association Between Baseline CAD and the Development of Epilepsy in the Longitudinal Cohort 1

As shown in **Supplemental Table 2**, during a median follow-up of 13.82 years, a total of 3,590 (0.72%) out of 496,921 participants developed new-onset epilepsy. Among these, 3,183 (0.68%) cases occurred in the non-CAD group, while 407 (1.52%) cases were observed in the CAD group. The Kaplan-Meier curve demonstrates a significantly higher cumulative incidence of epilepsy in the CAD group compared to the non-CAD group (log-rank *P* < 0.001, **Figure 2A**). Subsequently, we assessed the association between CAD and new-onset epilepsy (**Table 3**). After adjusting for all potential confounders, the HR for the CAD group was 1.32 (95% CI, 1.15–1.51, *P* < 0.001).

**Figure 2.**
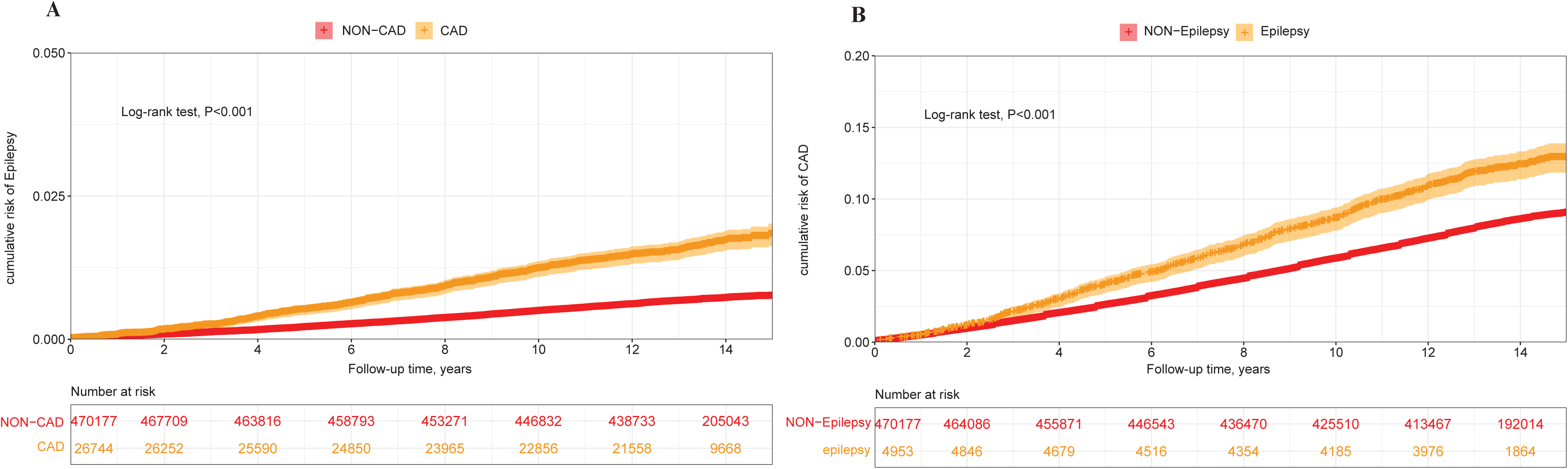
Cumulative risk of epilepsy in patients with CAD and cumulative risk of CAD in patients with epilepsy. (A) Cumulative risk of epilepsy in CAD participants and non-CAD participants. (B) Cumulative risk of CAD in epilepsy and non-epilepsy participants. Abbreviation: CAD, coronary artery disease.

**Table 3.** Association Between Baseline CAD and the Incidence of epilepsy in the Prospective Cohort Analysis.

| Groups | No. of<br>Participants | Events/Person-years | Model 1 |  | Model 2 |  | Model 3 |  |
| --- | --- | --- | --- | --- | --- | --- | --- | --- |
|  |  |  | HR (95% CI) | P-value | HR (95% CI) | P-value | HR (95% CI) | P-value |
| Non-CAD | 470177 | 3183/6721643 | 1 (reference) | - | 1 (reference) | - | 1 (reference) | - |
| CAD | 26744 | 407/354194 | 1.86 (1.67-2.06) | < 0.001 | 1.60 (1.42-1.82) | < 0.001 | 1.32 (1.15-1.51) | < 0.001 |

Model 2: adjusted for age, sex, BMI, ethnicity, alcohol drinking status, smoking status, physical activity, education, UK Biobank assessment centre, and Townsend deprivation index.
Model 3: adjusted for age, sex, BMI, ethnicity, alcohol drinking status, smoking status, physical activity, education, UK Biobank assessment centre, Townsend deprivation index, LDL cholesterol, total triglycerides, SBP, GLU, CRP, and comorbidity/medication history (hypertension, hyperthyroidism, diabetes, depression, cerebrovascular disease, hypertriglyceridemia, central nervous system inflammation, cholesterol-lowering drug, antihypertensive drug, insulin).
Abbreviation: CAD, coronary artery disease; BMI, body mass index; LDL, low-density lipoprotein; HDL, high-density lipoprotein; GLU, glucose; SBP, systolic blood pressure; CRP, C-reactive protein.

Furthermore, we divided participants into low and high genetic risk groups based on their median epilepsy PRS score and conducted a subgroup analysis. (**Figure 3A**). Interestingly, in the multivariate adjusted Cox regression model (n=481,922), we found that the association between CAD and new-onset epilepsy was significant only among participants with low genetic susceptibility to epilepsy (HR, 1.41, 95% CI, 1.17–1.70, *P* < 0.001).

**Figure 3.**
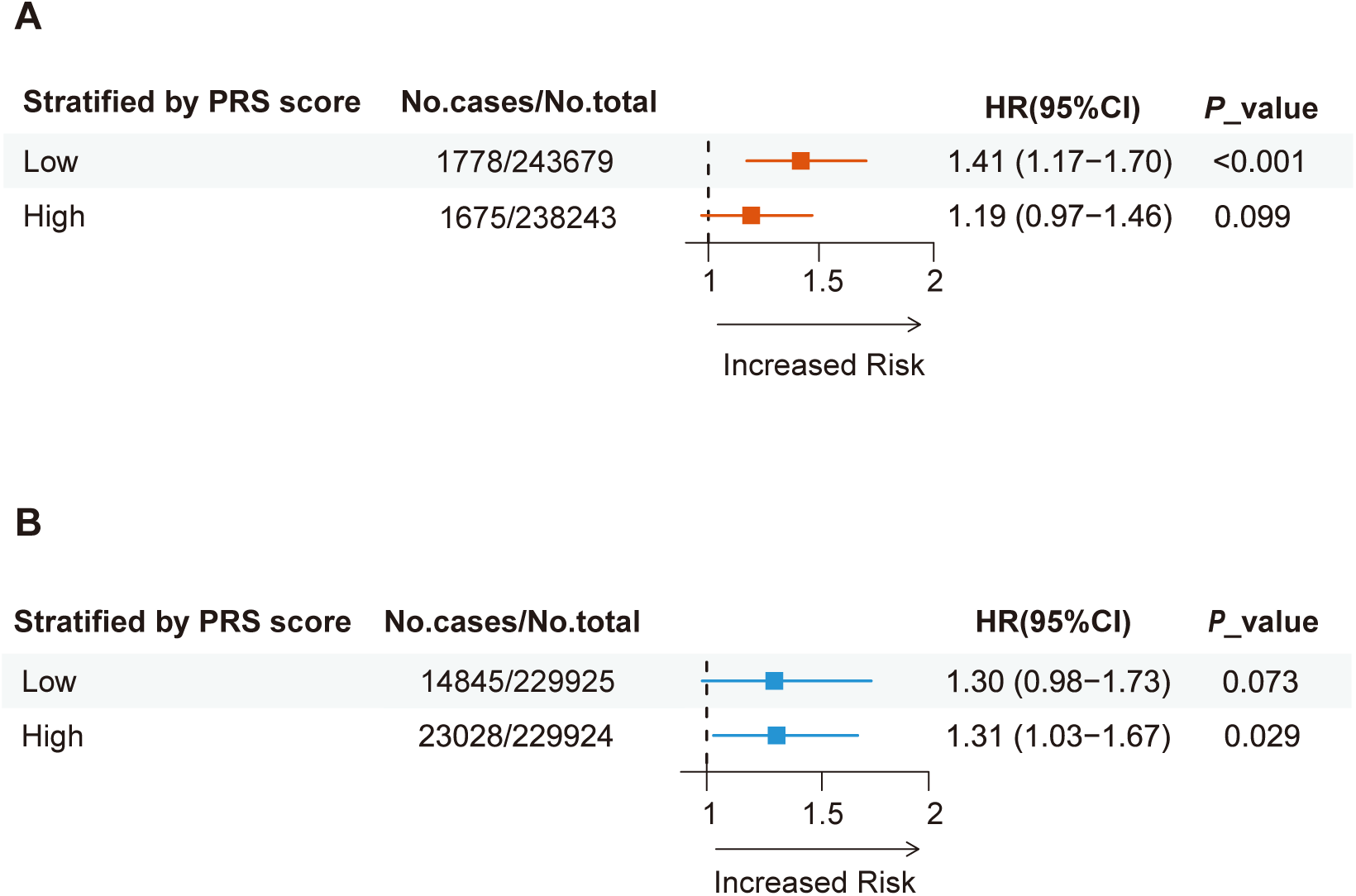
Genetic susceptibility-adjusted bidirectional analysis of epilepsy and CAD in the prospective cohort. (A) Association between CAD and incident epilepsy, stratified by or adjusted for genetic susceptibility to epilepsy. (B) Association between epilepsy and incident CAD, stratified by or adjusted for genetic susceptibility to CAD. Model 1: adjusted for age, sex, and ethnicity. Model 2: adjusted for age, sex, BMI, ethnicity, alcohol drinking status, smoking status, physical activity, education, and Townsend deprivation index. Model 3: adjusted age, sex, BMI, ethnicity, alcohol drinking status, smoking status, physical activity, education, Townsend deprivation index,total cholesterol, LDL cholesterol, HDL cholesterol, total triglycerides, glucose, systolic blood pressure, C-reactive protein, and Comorbidity/medication history. Abbreviation: CAD, coronary artery disease; BMI, body mass index; LDL, low density lipoprotein; HDL, high density lipoprotein.

Finally, we conducted subgroup analyses based on age, sex, BMI, ethnicity, smoking status, alcohol consumption, physical activity, and comorbidities (**Figure 4**). Notably, we found that the association remained significant regardless of age, sex, obesity status, physical activity levels, and among White participants, as well as in individuals with comorbidities. Additionally, we found an interaction between sex, smoking status, and the effect of CAD on epilepsy. (*P* for interaction < 0.05).

**Figure 4.**
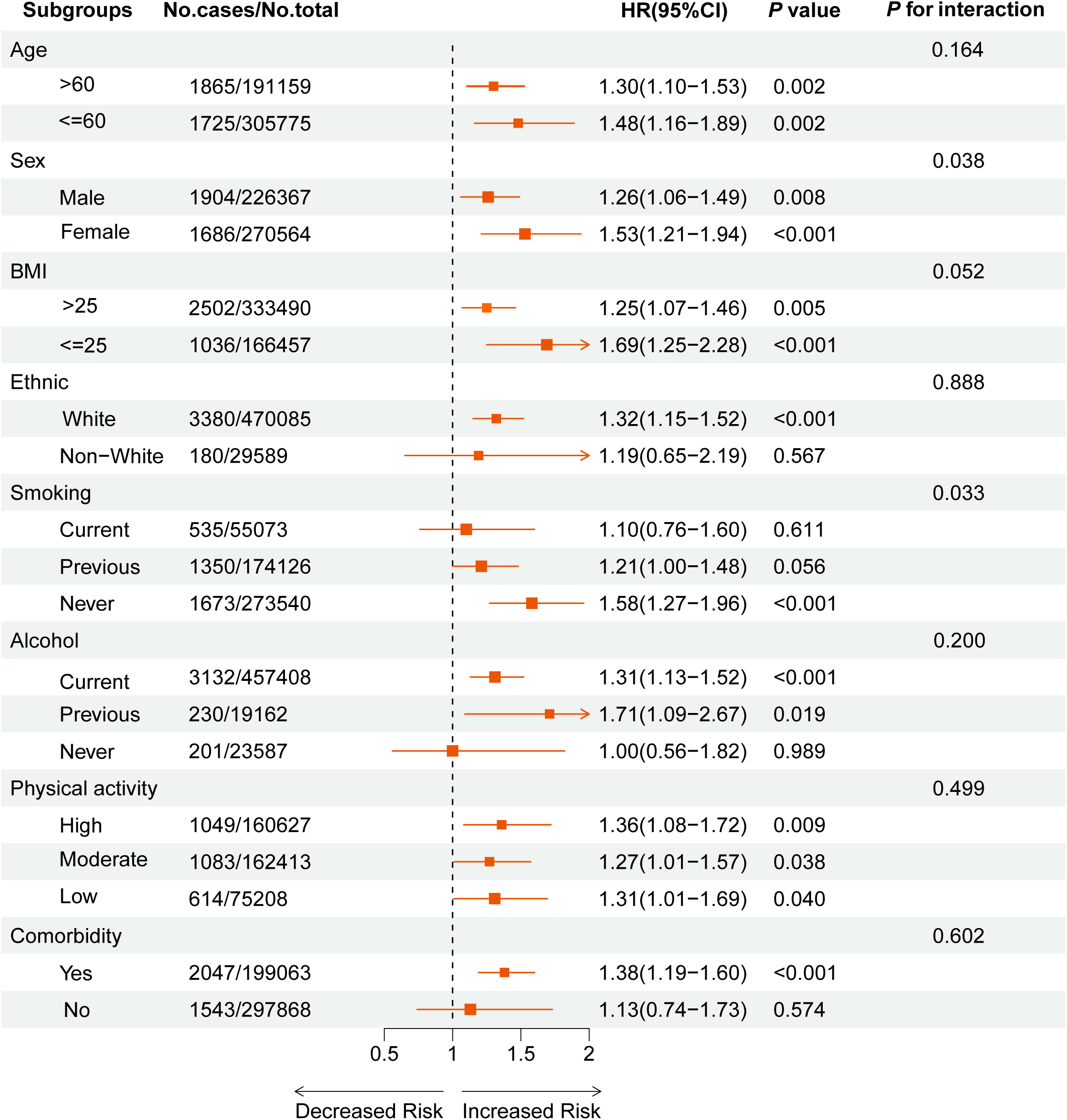
The relationship between CAD and the incidence of epilepsy, stratified by covariates. Model adjusted age, sex, BMI, ethnicity, alcohol drinking status, smoking status, physical activity, education, Townsend deprivation index, total cholesterol, LDL cholesterol, HDL cholesterol, total triglycerides, glucose, systolic blood pressure, C-reactive protein, and comorbidity/medication history. (Omit the corresponding grouping variable from the relevant subgroup analysis (e.g., do not adjust for the sex factor in the sex subgroup)). Abbreviation: CAD, coronary artery disease; BMI, body mass index; LDL, low density lipoprotein; HDL, high density lipoprotein.

### Association Between Baseline Epilepsy and the Development of CAD in the Longitudinal Cohort 2

In analyzing the risk of incident CAD associated with epilepsy, we followed a total of 475,130 participants for a median of 13.73 years. During this period, 576 (11.6%) epilepsy participants and 38,647 (8.2%) non-epilepsy individuals developed CAD (**Supplemental Table 3**). The Kaplan-Meier curve demonstrates a significant difference in the incidence of CAD between participants with and without epilepsy (log-rank *P* < 0.001, **Figure 2B**). We subsequently assessed the association between baseline and new-onset epilepsy. We found that participants with epilepsy showed a significantly increased risk of developing CAD (**Table 4**). After adjusting for all potential confounders, the HR for the epilepsy group was 1.31 (95% CI, 1.12–1.54, *P* < 0.001).

**Table 4.** Association Between Baseline epilepsy and the Incidence of CAD in the Prospective Cohort Analysis.

| Groups | No. of<br>Participants | Events/Person-years | Model 1 |  | Model 2 |  | Model 3 |  |
| --- | --- | --- | --- | --- | --- | --- | --- | --- |
|  |  |  | HR(95% CI) | P-value | HR(95% CI) | P-value | HR(95% CI) | P-value |
| Non-Epilepsy | 470177 | 38647/6499446 | 1 (reference) | - | 1 (reference) | - | 1 (reference) | - |
| Epilepsy | 4953 | 576/65143 | 1.52 (1.40-1.65) | < 0.001 | 1.28 (1.12-1.47) | < 0.001 | 1.31 (1.12-1.54) | < 0.001 |

Model 1: adjusted for age, sex, and ethnicity.
Model 2: adjusted for age, sex, BMI, ethnicity, alcohol drinking status, smoking status, physical activity, and education, UK Biobank assessment centre, and Townsend deprivation index.
Model 3: adjusted for age, sex, BMI, ethnicity, alcohol drinking status, smoking status, physical activity, education, UK Biobank assessment centre, Townsend deprivation index, LDL cholesterol, total triglycerides, SBP, GLU, CRP, and comorbidity/medication history (hypertension, hyperthyroidism, diabetes, depression, cerebrovascular disease, hypertriglyceridemia, CVD family history, cholesterol-lowering drug, antihypertensive drug, insulin).
Abbreviation: CAD, coronary artery disease; BMI, body mass index; LDL, low-density lipoprotein; HDL, high-density lipoprotein; GLU, glucose; CRP, C-reactive protein; CVD, cardiovascular disease.

Moreover, subgroup analysis stratified by the median PRS (n=459,849) for CAD revealed that although the association between epilepsy and new-onset CAD was significant only in participants with high genetic risk, there was no significant difference in risk between low genetic risk and high genetic risk participants. The HRs were 1.30 (95% CI, 0.98–1.73, *P* = 0.073) for low genetic risk participants and 1.31 (95% CI, 1.03–1.67, *P* = 0.029) for high genetic risk participants (**Figure 3B**). Furthermore, we conducted subgroup analyses based on age, sex, BMI, ethnicity, smoking status, alcohol consumption, physical activity, and comorbidities (**Figure 5**). We found that the association between epilepsy and new-onset CAD remained significant regardless of age, BMI, and among male participants, white participants, current smokers, current drinkers, and those with comorbidities. Additionally, we identified an interaction between BMI and the effect of epilepsy on CAD (*P* for interaction < 0.05).

**Figure 5.**
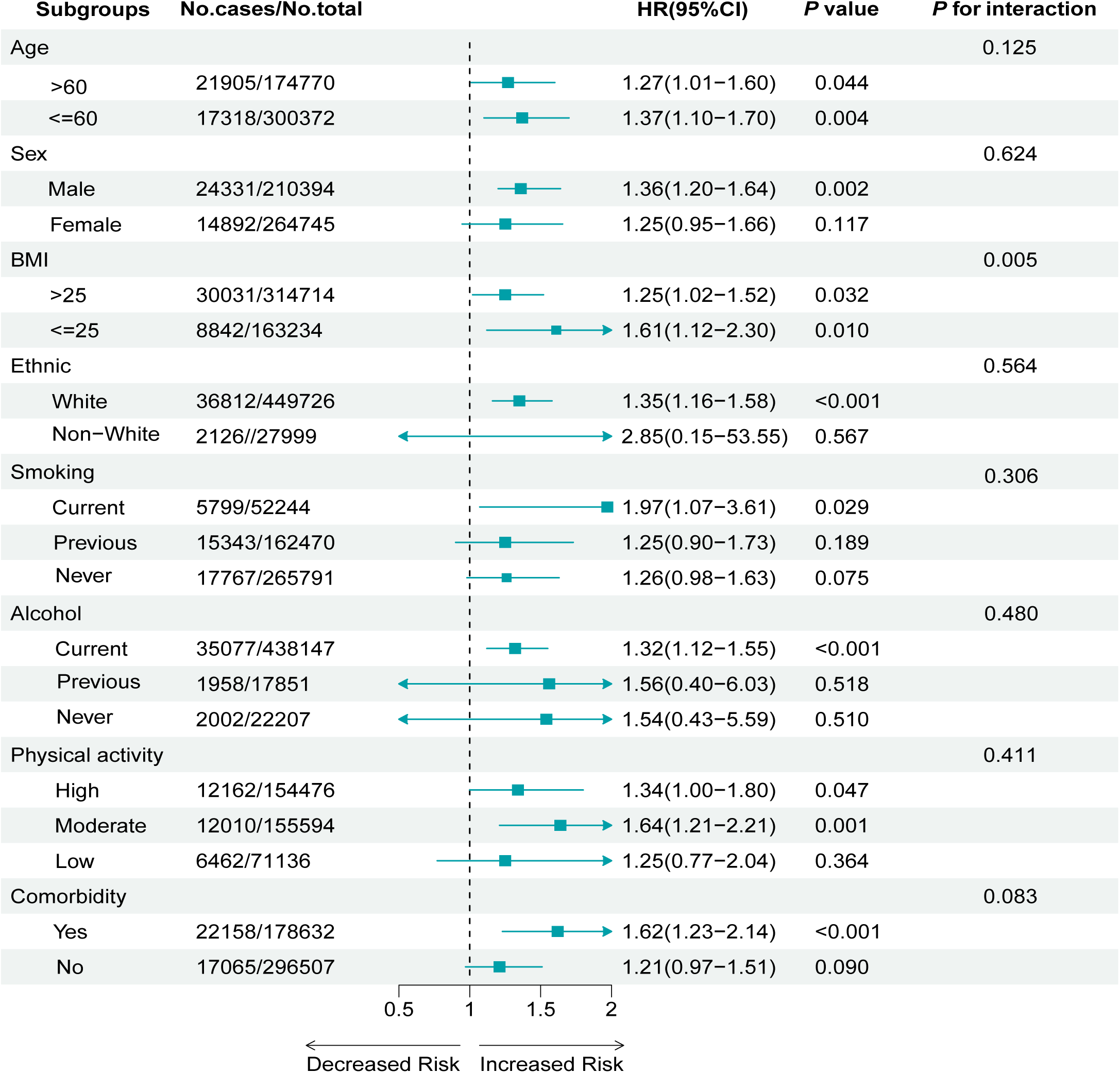
The relationship between epilepsy and the incidence of CAD, stratified by covariates. Model adjusted age, sex, BMI, ethnicity, alcohol drinking status, smoking status, physical activity, education, Townsend deprivation index, total cholesterol, LDL cholesterol, HDL cholesterol, total triglycerides, glucose, systolic blood pressure, C-reactive protein, and comorbidity/medication history. (Omit the corresponding grouping variable from the relevant subgroup analysis (e.g., do not adjust for the sex factor in the sex subgroup)). Abbreviation: CAD, coronary artery disease; BMI, body mass index; LDL, low density lipoprotein; HDL, high density lipoprotein.

### Sensitivity analysis

Several additional sensitivity analyses were performed to evaluate the robustness of our findings, with results detailed in **Supplementary Tables 4-7**. These analyses included excluding participants with a follow-up duration of less than two years, restricting the primary analysis to participants with complete major covariate data, applying Fine-Gray competing risk models, and using a mixed-effects model to adjust for institutional factors. The results of all sensitivity analyses were generally consistent with the main conclusion.

## Discussion

This large cross-sectional and prospective cohort study provides the first evidence of a bidirectional association between CAD and epilepsy. First, cross-sectional studies have shown a clear association between CAD and epilepsy. Additionally, longitudinal cohort studies have demonstrated that individuals with CAD are at a higher risk of new-onset epilepsy than those without CAD. Likewise, individuals with epilepsy are more likely to experience incident CAD compared to those without epilepsy. Furthermore, our findings indicate that the association between CAD and new-onset epilepsy was significant only in participants with low genetic susceptibility. In contrast, the association between epilepsy and new-onset CAD was significant only in participants with high genetic susceptibility. Finally, the findings were consistent across subgroups stratified by age, sex, and BMI. Our findings not only deepen our understanding of the mechanisms underlying the bidirectional relationship between these two diseases, but also have the potential to inform more integrated preventive and therapeutic strategies in clinical practice.

Our study on the bidirectional relationship between CAD and epilepsy extends previous research findings. Cross-sectional studies in the U.S. population have shown that 21% of adults with epilepsy also reported a history of heart disease, 9% higher than the 12% observed in adults without epilepsy. Additionally, adults with a history of heart disease were nearly twice as likely to report a history of epilepsy compared to those without a history of epilepsy.^24^ In addition to the cross-sectional analysis of the prevalence and association between epilepsy and CAD, our study also considered the temporal sequence of the two conditions. We further used two longitudinal cohorts to explore the relationship between baseline CAD and incident epilepsy, as well as between baseline epilepsy and incident CAD. Previous cohort studies from the United States and the United Kingdom have reported findings consistent with ours, indicating a higher prevalence of cardiovascular disease among individuals with epilepsy and a greater incidence of heart disease during follow-up.^26–28^ Building upon these findings, we further demonstrated that individuals with CAD are at a higher risk of new-onset epilepsy than those without CAD. Likewise, individuals with epilepsy are at a higher risk of incident CAD than those without epilepsy. Additionally, we investigated the potential influence of genetic susceptibility on the association between the two conditions. Our study adds to the existing literature and highlights that we should take a holistic approach to managing both disorders and provide integrated care that includes health promotion, disease prevention, diagnosis, treatment, and prognosis to reduce the burden of these two co-existing disorders.

Our study also explored the relationship between CAD and epilepsy across different subgroups. We found that the association remained statistically significant regardless of age or BMI level. This may be attributed to shared risk factors and the presence of brain-heart interactions, which are observed across different age and BMI groups.^51^ In the sex-stratified analysis, the association between baseline epilepsy and incident CAD was statistically significant in males but not in females. This difference may be related to sex-specific baseline cardiovascular risk. Males generally have a higher risk of CAD at a younger age, which may amplify the observable effect of epilepsy on CAD. In contrast, premenopausal females are protected by estrogen, which lowers their cardiovascular risk and may mask the influence of epilepsy.^52^ Furthermore, we observed that the bidirectional association between epilepsy and CAD was significant among individuals with comorbidities, but not among those without. This may be due to the role of comorbidities as ‘amplifiers’ of risk or as mediating factors that enhance the interaction between epilepsy and CAD through mechanisms such as chronic inflammation, endothelial dysfunction, and pharmacological interactions.^53–54^

Interestingly, the association between baseline CAD and incident epilepsy was statistically significant only among individuals with low genetic susceptibility. One possible explanation is that individuals with a high genetic risk for epilepsy may possess more responsive neuroprotective feedback mechanisms. For example, enhanced regulatory functions of calcium channel-related genes may attenuate the impact of exogenous risk factors such as CAD on brain function, thereby masking their pro-epileptogenic effects.^61^ In contrast, the effect of baseline epilepsy on incident CAD was significant only among individuals with high genetic susceptibility. Existing studies have indicated that epilepsy and CAD may share common risk factors and partially overlapping genetic risk loci, such as those associated with chronic inflammation, oxidative stress, and ion channel dysfunction.^62–65^ However, to date, no large-scale genome-wide association studies (GWAS) have systematically examined the shared genetic variants between these two conditions. Future studies are needed to investigate this potential genetic relationship.

One of the mechanisms explaining the bidirectional relationship between CAD and epilepsy may be the presence of common risk factors. These include metabolic syndrome, medications, lifestyle, genetic predisposition, and other factors.^19,49,62–64^ Moreover, epilepsy patients may be at higher risk for CAD due to several interrelated mechanisms. One hypothesis is that seizure-induced catecholamine release causes repetitive myocardial injury (’epileptic heart’), resulting in chronic myocardial and coronary artery damage, which promotes myocardial fibrosis, accelerated atherosclerosis, and cardiac dysfunction^37–39^ Additionally, certain antiepileptic drugs (AEDs) have been shown to adversely affect lipid metabolism and blood glucose levels, thereby increasing the risk of CAD.^55–56^ Furthermore, seizures often trigger a stress response with elevated glucocorticoids, which may promote atherosclerosis by impairing endothelial function and reducing vasodilation, thereby increasing CAD risk.^57^ Apart from the abovementioned effects between CAD and epilepsy, patients with CAD may have an increased risk of epilepsy due to several interrelated factors. CAD often reduces systemic blood flow, leading to cerebral ischemia and subsequent hypoxia. These conditions can alter brain electrophysiology, increasing susceptibility to epileptic activity.^58–59^ Moreover, CAD is frequently accompanied by chronic inflammation and oxidative stress, both of which can contribute to neuronal damage and disrupt normal electrical activity in the brain, further elevating the risk of epilepsy.^60^

Our study has several limitations. First, the observed association between epilepsy and CAD is based on evidence from cross-sectional and cohort studies, which cannot establish a causal relationship. Second, although our observational analysis adjusted for a wide range of potential confounders, residual confounding cannot be entirely excluded, and correction for the potential effects of antiepileptic drugs on CAD was not possible due to missing data. Third, as most participants in the UK Biobank were of UK descent, our findings may not be generalizable to other ethnic populations.

### Conclusion

This study is the first to systematically reveal the bidirectional temporal relationship between CAD and epilepsy in a large-scale population, while also exploring the heterogeneity of this association across different subpopulations and its potential modification by genetic susceptibility. Our findings suggest that for these two highly comorbid conditions, interdisciplinary collaboration and integrated disease management should be strengthened to enable more effective health interventions and improved clinical outcomes.

## Declarations

### Ethics approval and consent to participate

The UK Biobank was approved by the North West Multi-Centre Research Ethics Committee. All participants provided written and informed consent for data collection, analysis, and record linkage. This study was performed under UK Biobank application number 77195.

### Consent for publication

Not applicable.

### Availability of data and materials

This research has been conducted using the UK Biobank resource under application number [77195]. The data are not publicly available but can be accessed by bona fide researchers upon application to the UK Biobank (https://www.ukbiobank.ac.uk/).

### Competing interests

The authors declare that they have no competing interests.

## Funding

This work was supported by the National Science Foundation of China (82270390, JCRCFZ-2022-006), the Hubei Province Innovation Platform Construction Project (20204201117303072238), and the Jiangxi Province Thousand Talents Project (jxsq2023101002).

## Authors’ contributions

Li Zhang: Conceptualization, Methodology, Investigation, Visualization, Writing – original draft, Writing – review & editing, Supervision. Weifang Liu: Conceptualization, Investigation, Data curation, Visualization, Writing – original draft, Writing – review & editing. Yilu Xue: Methodology, Data curation, Validation. Liwen Wang: Methodology, Data curation, Validation. Cong Gong: Methodology, Data curation, Validation. Zhi-Gang She: Supervision, Project administration, Funding acquisition. Hongliang Li: Conceptualization, Supervision, Methodology, Resources, Writing – review & editing, Funding acquisition, Project administration.

## Data Availability

The data that support the findings of this study are available from the UK Biobank (application number: 77195), but restrictions apply to the availability of these data, which were used under license for the current study and are not publicly available. Data are, however, available from the authors upon reasonable request and with permission from the UK Biobank.

## Acknowledgements

We sincerely thank the participants and staff of the UK Biobank for their invaluable contributions to this research. This study was conducted using data from the UK Biobank under application number 77195, with ethical approval granted by the North West Multi-centre Research Ethics Committee (REC reference: 11/NW/03820).

## List of abbreviations

CAD: Coronary artery disease
CI: Confidence interval
HR: Hazard ratio
OR: Odds ratio
PRS: Polygenic risk score
UKB: UK Biobank
BMI: Body mass index
GWAS: Genome-wide association study
SNP: Single nucleotide polymorphism
LDL: Low density lipoprotein
HDL: high density lipoprotein
DBP: diastolic blood pressure
SBP: systolic blood pressure
CRP: C-reactive protein
GLU: glucose
CVD: cardiovascular disease

## Notes

### Competing Interest Statement

The authors have declared no competing interest.

### Clinical Trial

This study is a prospective observational cohort study using data from the UK Biobank. It is not a clinical trial and therefore was not registered in a trial registry such as ClinicalTrials.gov.

### Author Declarations

This study was conducted using data from the UK Biobank under application number 77195. The UK Biobank has ethical approval from the North West Multi-Centre Research Ethics Committee (MREC), and all participants provided informed consent. The use of these data for this research was approved by the UK Biobank Ethics and Governance Council.

